# Topological Alterations in Brain Functional Connectivity between ASD and Typically Developing Individuals: A Graph-Theoretical Analysis using Multi-Site Resting-State fMRI Data

**DOI:** 10.64898/2026.01.27.26344914

**Authors:** Talha Imtiaz Baig, Hongzhou Wu, XiYang Li, Junlin Jing, Bharat B. Biswal, Benjamin Klugah-Brown

## Abstract

Autism spectrum disorder (ASD) is increasingly conceptualized as a disorder of large-scale functional brain network organization rather than isolated regional abnormalities. Graph-theoretical analysis provides a principled framework for characterizing such distributed network reconfiguration. Here, we investigated global, nodal, and system-level functional network topology in ASD using a large, multi-site resting-state fMRI dataset. Resting-state fMRI data from 996 participants (428 ASD, 568 healthy controls) were obtained from the ABIDE I and II data repositories. Whole-brain weighted resting state functional networks were constructed using Pearson correlation. To improve robustness and reduce threshold-selection bias, graph-theoretical metrics were computed across a range of network sparsity thresholds and summarized using an area-under-the-curve (AUC) approach. At the global level, ASD was associated with reduced assortativity, and local efficiency, along with altered normalized characteristic path length (λ), indicating local information processing and subtle deviations in network integration relative to an optimal small-world topology. Nodal analyses revealed non-random, region-specific alterations predominantly affecting higher-order associative systems. Increased nodal centrality and hub-like properties were observed in frontal and parietal regions within the frontoparietal control and dorsal attention networks, whereas reduced nodal efficiency and centrality were primarily localized to limbic and anterior temporal regions, including the temporal pole. System-level analyses, controlling for age, sex, and acquisition site, further demonstrated network-specific topological reorganization across multiple functional systems. Clinical correlation analyses identified modest but significant associations between nodal topology and core ASD symptom severity, particularly within default mode, limbic, and attention networks. Together, these findings indicate that ASD is characterized by subtle yet reproducible multi-scale reorganization of functional brain network topology, supporting a systems-level account of ASD neurobiology and highlighting the clinical relevance of large-scale network architecture.

## 1 Introduction

Autism spectrum disorder (ASD) is a neurodevelopmental condition that emerges early in life and is characterized by persistent difficulties in social communication, restricted interests, and repetitive behaviors[1]. Beyond focal regional abnormalities, converging evidence indicates that ASD is associated with widespread alterations in the organization of large-scale brain networks, affecting both local information processing and long-range communication between distributed brain regions [2-4]. A central feature of ASD neurobiology is altered functional connectivity (FC), reflecting atypical coordination of neural activity across the brain [5]. Understanding how FC patterns are reorganized in ASD is critical for clarifying the neural mechanisms underlying symptom heterogeneity and for linking brain-level alterations to clinical manifestations [6-8].

ASD is a heterogeneous condition, with substantial inter-individual variability in symptom severity, cognitive functioning, and developmental trajectory [9]. Multiple functional brain systems, including those supporting social cognition, attention, executive control, sensory processing, and emotion regulation, have been implicated in core ASD symptoms[10-12]. Consequently, a major goal of contemporary ASD research is to characterize how large-scale brain networks are organized and how deviations from typical network architecture relate to behavioral and clinical phenotypes. The increasing prevalence of ASD reported worldwide [13, 14] further underscores the importance of identifying robust and generalizable neurobiological markers.

Resting-state functional magnetic resonance imaging (rs-fMRI) provides a powerful, task-free approach for probing intrinsic brain organization by quantifying temporal correlations in spontaneous blood oxygenation level-dependent (BOLD) signals across spatially distributed regions of interest [15, 16]. Numerous rs-fMRI studies have reported atypical connectivity in ASD across regions implicated in social cognition, emotion, and executive function, including the prefrontal cortex, temporal lobe, amygdala, cerebellum, and insula [17-19].

Graph theoretical functional brain networks are commonly constructed by estimating pairwise functional connectivity between brain regions and organizing these relationships into network representations [20]. Canonical resting-state networks, such as the default mode network (DMN), frontoparietal control network (FPN), dorsal and ventral attention networks (DAN/VAN), sensorimotor network (SMN), and visual network (VSN), have been consistently identified and linked to distinct cognitive and behavioral functions [11, 12]. Several of these networks, particularly the DMN, attention networks, and control networks, have been repeatedly implicated in ASD-related impairments in social cognition, attentional control, and executive functioning. While traditional FC analyses have yielded valuable insights, they typically rely on pairwise associations and therefore provide limited information about the global and mesoscale organization of brain networks.

Graph theory offers a principled mathematical framework for modeling the brain as a complex network composed of nodes (brain regions) and edges (functional connections), enabling quantitative characterization of network topology across multiple spatial scales [21]. Graph-theoretical metrics capture complementary aspects of network organization, including integration (e.g., global efficiency, characteristic path length), segregation (e.g., clustering coefficient, local efficiency), and centrality or hubness (e.g., degree and betweenness centrality) [22-25]. These measures have been widely applied to investigate large-scale network alterations across neuropsychiatric and neurodevelopmental disorders, including ASD, schizophrenia, bipolar disorder, and ADHD [26]. Importantly, graph theory allows researchers to move beyond isolated connectivity differences and instead examine how brain-wide communication architecture is reorganized.

Despite substantial progress, prior FC and graph-theoretical studies in ASD have produced inconsistent and sometimes contradictory findings, with reports of hypo-connectivity, hyper-connectivity, or mixed alterations depending on analytic choices [6, 21, 25]. Such variability likely reflects differences in sample size, age range, preprocessing pipelines, parcellation schemes, thresholding strategies, and limited generalizability of single-site datasets. Moreover, many studies have focused on relatively small or homogeneous samples, limiting the ability to identify network features that are robust across sites and populations. These challenges highlight the need for large-scale, multi-site investigations employing standardized and methodologically robust graph-theoretical approaches.

The Autism Brain Imaging Data Exchange (ABIDE) provides an opportunity to address these limitations by enabling large-sample, multi-site analyses of rs-fMRI data in ASD. When combined with proportional thresholding and area-under-the-curve (AUC) integration across sparsity levels, graph-theoretical analysis can reduce threshold-selection bias and improve the stability and reproducibility of network metrics. Such approaches allow for systematic characterization of global, nodal, and system-level topological differences between ASD and typically developing individuals.

Building on prior multi-site work that focused on component-level functional connectivity and classification using spatially constrained independent component analysis [27, 28], the present study extends this line of research by examining whether ASD is associated with systematic reorganization of whole-brain functional network topology. Specifically, we investigate global network properties, regional nodal characteristics, and network-wise (system-level) topology using weighted graph-theoretical analysis of rs-fMRI data from ABIDE I and II. We hypothesize that ASD is characterized by disrupted balance between network segregation and integration, reflected by deviations from an optimal small-world topology, reduced local efficiency, and redistribution of hub architecture within key functional systems, particularly the default mode, frontoparietal control, attention, and limbic networks. By leveraging a large, heterogeneous multi-site dataset and robust graph-theoretical methodology, this study aims to identify reproducible topological features of functional brain networks that distinguish individuals with ASD from healthy controls and clarify their potential clinical relevance.

## 2 Methods and Materials

This section describes the datasets, preprocessing procedures, network construction methods, graph-theoretical analyses, and statistical approaches used to examine large-scale functional brain network organization in ASD.

### 2.1 Dataset and Participants

Resting-state functional and structural MRI data were obtained from the Autism Brain Imaging Data Exchange (ABIDE I and ABIDE II) repositories (http://fcon_1000.projects.nitrc.org/indi/abide/) [29, 30]. The combined ABIDE dataset includes imaging and phenotypic data from 2,264 participants recruited across 38 international sites, comprising 1,083 individuals diagnosed with ASD and 1,181 typically developing healthy controls (HCs). ASD diagnoses were established at each contributing site using standardized clinical assessments consistent with established diagnostic criteria, including the Autism Diagnostic Observation Schedule (ADOS) [31] and Autism Diagnostic Interview or equivalent diagnostic instruments [32, 33]. Demographic and cognitive variables, including age, sex, full-scale IQ (FIQ), verbal IQ (VIQ), and performance IQ (PIQ), when available, were assessed to characterize group differences and sample composition. All ABIDE datasets were collected under local institutional ethics approvals with informed consent/assent, and were shared publicly in de-identified form by the contributing sites.

### 2.2 fMRI Data Preprocessing

Preprocessing was performed using DPARSF as implemented in “DPARSFA” embedded setting with DPABI toolkit (DPARSF; http://www.rfmri.org) [34, 35], running in MATLAB (R2022b). Preprocessing followed standard and widely adopted procedures for multi-site resting-state fMRI (rs-fMRI) analysis: the first 10 volumes were removed to allow for scanner signal stabilization; slice-timing correction was applied [36]; rigid-body realignment corrected for head motion [37]; functional images were co-registered to individual T1-weighted structural images; tissue segmentation and nonlinear normalization were carried out using DARTEL. Given ongoing debate regarding GSR, results should be interpreted within a no-GSR preprocessing framework, particularly regarding its impact on functional connectivity estimates and its potential to introduce spurious negative correlations [38-40]; spatial normalization to MNI space was conducted with a voxel resolution of 3□×□3□×□3□mm^3^ using DARTEL [41, 42]; nuisance covariates, including cerebrospinal fluid signals, were regressed out to reduce low-frequency drift and high-frequency components commonly attributed to physiological and scanner noise[43]; and spatial smoothing was performed using a 6-mm full-width at half-maximum (FWHM) Gaussian kernel [44]. These preprocessing steps were applied uniformly across all sites to ensure methodological consistency.

### 2.3 Quality Control

To enhance data quality and ensure reliable group comparisons, several exclusion criteria were applied. Sites with minimal ASD representation (e.g., CMU from ABIDE I) or those containing only ASD participants (e.g., KUL and NYU2 from ABIDE II) were excluded to allow for meaningful between-group analyses. Individual participants were removed if they exhibited excessive head motion, defined as translation >2 mm, rotation >2°, and FD was computed from realignment parameters (definition as implemented in DPABI), and participants with mean FD > 0.2 mm were excluded[45, 46], had missing phenotypic data, displayed poor image quality, or provided insufficient time points. Total 432 participants were excluded for these reasons. After this initial quality control, data from 1,750 participants across 35 sites remained. To further enhance statistical robustness and group balance, only sites contributing more than 50 participants with both ASD and HC individuals were retained. This criterion was used to improve stability of site-specific variance estimates and reduce small-site sampling noise. The final analytical sample comprised 996 participants from 11 sites: 428 with ASD and 568 HCs, aged 5 to 64 years. Reflecting the typical demographic profile of ASD cohorts, the sample included a higher proportion of male participants. This curated dataset formed the foundation for all subsequent analyses.

### 2.4 ROI Time Series Extraction

Regions of interest (ROIs) were defined using the Schaefer 2018 atlas, which includes cortical parcels only, therefore the present research focused on the organization of large scale cortical 17 functional networks [47]. This atlas integrates functional connectivity gradients with the Yeo intrinsic network framework, offering high reproducibility and a balance between system-level interpretability and spatial resolution suitable for nodal analyses [10]. For each participant, preprocessed 4D resting-state fMRI data were imported into DPABI, and ROI time series extraction was performed in MNI space after spatial normalization by mapping voxels to their corresponding Schaefer parcels. The mean BOLD time series for each ROI was then extracted by averaging the voxel-wise signals within each parcel, yielding a 200 × T time series matrix per participant, where T represents the number of time points. This extraction method aligns with established practices in graph-theoretical fMRI research employing similar or alternative parcellation schemes [48, 49].

### 2.5 Functional Connectivity Matrix Construction

Functional connectivity (FC) between all pairs of regions of interest (ROIs) was quantified using the Pearson correlation coefficient [50]. For ROIs *i* and *j*, with respective BOLD time series *x*_*i*_(*t*)and *x*_*j*_(*t*), FC was computed as:

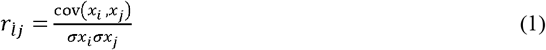

To improve the normality of the correlation distribution, all r values were subsequently transformed using Fisher’s r-to-z transformation. This yielded a symmetric 200 × 200 FC matrix for each participant. Both positive and negative correlations were retained in the functional connectivity metrics in accordance with standard practice in resting-state fMRI analysis. However, only positive weights were included for graph theoretical network construction and metric computation to ensure interpretability and consistency with efficiency related and shortest path-based metrics.

### 2.6 Graph-Theoretical Network Constructions

Graph-theoretical analyses were conducted using the GRETNA toolbox (version 2.0) [51]. To ensure consistent network density across participants and avoid biases due to inter-individual differences in connection strength, weighted brain networks were constructed from the FC matrices using proportional thresholding [52]. Sparsity levels ranged from 0.05 to 0.50 in steps of 0.05, while lower sparsity (<0.05) may result in disconnected networks and higher sparsity (>0.05) can mask topological differences and the selected step size (0.05) provide enough resolution to capture topological variations to maintain computational efficiency [53] where sparsity (*S*) is defined as:

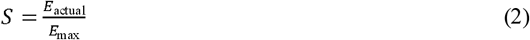

with *E*_actual_ representing the number of retained edges and *E*_max_the total possible edges in a fully connected network. Networks were maintained as weighted to preserve the continuous nature of functional connectivity and minimize information loss associated with binarization [54]. Consistent with common practice in FC-based graph analysis, only positive correlations were included in the final graph construction. For each sparsity level, 100 random surrogate networks were generated to normalize global metrics relative to a null random graph model.

#### 2.6.2 Graph Measures

Graph-theoretical metrics were computed at three levels, global, nodal, and system, to capture complementary aspects of brain network organization [55]. Global measures included assortativity (*r*), global efficiency (*E*g), small-worldness (σ), global synchronization (*R*), and global hierarchy (β), with mathematical formulations following established conventions [45, 56-59]. Nodal measures characterized the functional role of individual regions and included clustering coefficient (CC), degree centrality (DC), betweenness centrality (BC), nodal efficiency (*Ei*), nodal local efficiency (*Eloc, i*), and shortest path length (*L*i) [60-63]. All these graph measures were selected to identify similar and biological meaningful dimensions of large-scale brain related to ASD. Global metrics evaluate whole brain information transfer efficiency, network consistency and small-world properties, while nodal metrics define the topological importance of altered hubs and regions specific network configuration [64, 65]. System-level measures were derived by aggregating nodal metrics within each of the 17 Yeo functional networks. Network segregation was defined as:

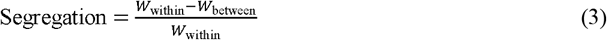

where *W*_within_and *W*_between_denote average within- and between-network connectivity strengths, respectively. System-level segregation was computed on the threshold, positive-weighted graphs at each sparsity level and summarized via area under the curve (AUC) [66]. Metrics were computed independently at each sparsity level and then summarized as a single AUC value per metric, providing a robust, threshold-independent scalar estimate. Threshold-specific results are reported in the Supplementary Materials.

### 2.7 Normality Check

Normality assessment [67] took place analytically instead of implicitly due to the large sample size. Distributional properties of global and nodal graph metrics were evaluated using visual inspection (quantile–quantile plots) to lead the selection of suitable statistical tests.

### 2.8 Statistical Analysis

Group differences between ASD and typically developing (HC) participants were assessed using two-tailed parametric two-sample *t*-tests for normally distributed metrics and Mann–Whitney *U* tests for non-normally distributed metrics (α = 0.05). Effect sizes were reported as Cohen’s *d* for parametric comparisons [68] and rank-biserial correlation for non-parametric tests [69]. To account for multiple comparisons across global, nodal, and system-level metrics, the Benjamini–Hochberg false discovery rate (FDR) procedure was applied with a significance threshold of *q* < 0.05 [70]. Clinical significance was identified between standardized ASD symptom severity measures (ADOS, ADI-R, and SRS) and nodal graph theoretical metrics by using Spearman rank correlation analysis. To reduce the cost of multiple comparisons, correlation analysis was limited to regions demonstrating convergent group level alterations across multiple nodal metrics. Analysis of covariance (ANCOVA) approach was used to evaluate group differences for system-level (network-wise) analysis. Age, sex and acquisition sites were included as covariates that adjusted for multi-site and demographic variability. AUC-based network metrics were compared between groups using covariate adjusted estimated marginal means.

## 3 Results

### 3.1 Demographic and Clinical Characteristics

Demographic and cognitive characteristics of the final sample (*N* = 996) are summarized in Table 1, with values reported as mean ± standard deviation unless otherwise noted. Group comparisons were conducted using chi-square tests for sex distribution and two-sample *t*-tests for age and IQ measures. The ASD and HC groups were well-matched in age and sex distribution, with no statistically significant differences observed (all *p* > 0.05). However, the ASD group showed significantly lower scores on full-scale IQ, verbal IQ, and performance IQ compared to HCs (all *p* < 0.001), a pattern consistent with findings from prior studies using ABIDE data. In order to avoid over controlling for variance intrinsically associated with the disorder, IQ was excluded as covariate in the main graph theoretical analysis because of well-established similarity between ASD diagnosis and IQ in large multi-site datasets. However, age, sex and acquisition sites were included as covariates where appropriate.

**Table 1:** Demographic and clinical characteristics of participants across sites.

| Variables | ASD (Mean ± SD) | HC (Mean ± SD) | Statistical test | p-value |
| --- | --- | --- | --- | --- |

| N | 428 | 568 | — | — |
| --- | --- | --- | --- | --- |
| **Sex (M/F)** | 373/55 | 418/150 | X<sup>2</sup> | 1.0000 |
| **Age (Years)** | 15.58 ± 9.06 | 14.85 ± 8.43 | t-test | 0.1945 |
| **Full Scale IQ** | 105.34 ± 16.46 | 113.46 ± 12.49 | t-test | <0.001 |
| **Verbal IQ** | 104.96 ± 17.84 | 114.75 ± 13.27 | t-test | <0.001 |
| **Performance IQ** | 104.97 ± 17.37 | 109.77 ± 13.26 | t-test | <0.001 |

Clinical profiles of the ASD cohort are detailed in Table 2, including standardized diagnostic and behavioral metrics such as Autism Diagnostic Observation Schedule (ADOS), Autism Diagnostic Interview-Revised (ADI-R), and Social Responsiveness Scale (SRS) scores.

**Table 2:** Clinical characteristics of ASD participants.

| Measures | Mean ± SD | N |
| --- | --- | --- |
| ADOS G Total | 11.34 ± 3.93 | 302 |
| ADOS G Social | 7.63 ± 2.72 | 223 |
| ADOS G Communication | 3.35 ± 1.52 | 223 |
| ADOS 2 Total | 12.60 ± 4.62 | 230 |
| ADOS 2 Severity Total | 6.95 ± 2.05 | 266 |
| SRS Total T | 76.75 ± 13.48 | 191 |
| SRS Awareness T | 62.09 ± 23.15 | 218 |
| SRS Cognition T | 64.49 ± 22.39 | 218 |
| SRS Communication T | 68.96 ± 21.01 | 218 |
| ADI-R Social Total A | 19.33 ± 5.33 | 318 |
| ADI-R RRB Total C | 5.80 ± 2.52 | 319 |

ASD cohort spans a broad range of symptom severity across core diagnostic domains, including social communication difficulties and repetitive behaviors. Site-specific related information is provided in Supplementary Tables S1 and S2. An overview of the complete analytical pipeline is illustrated in Figure 1.

**Figure 1.**
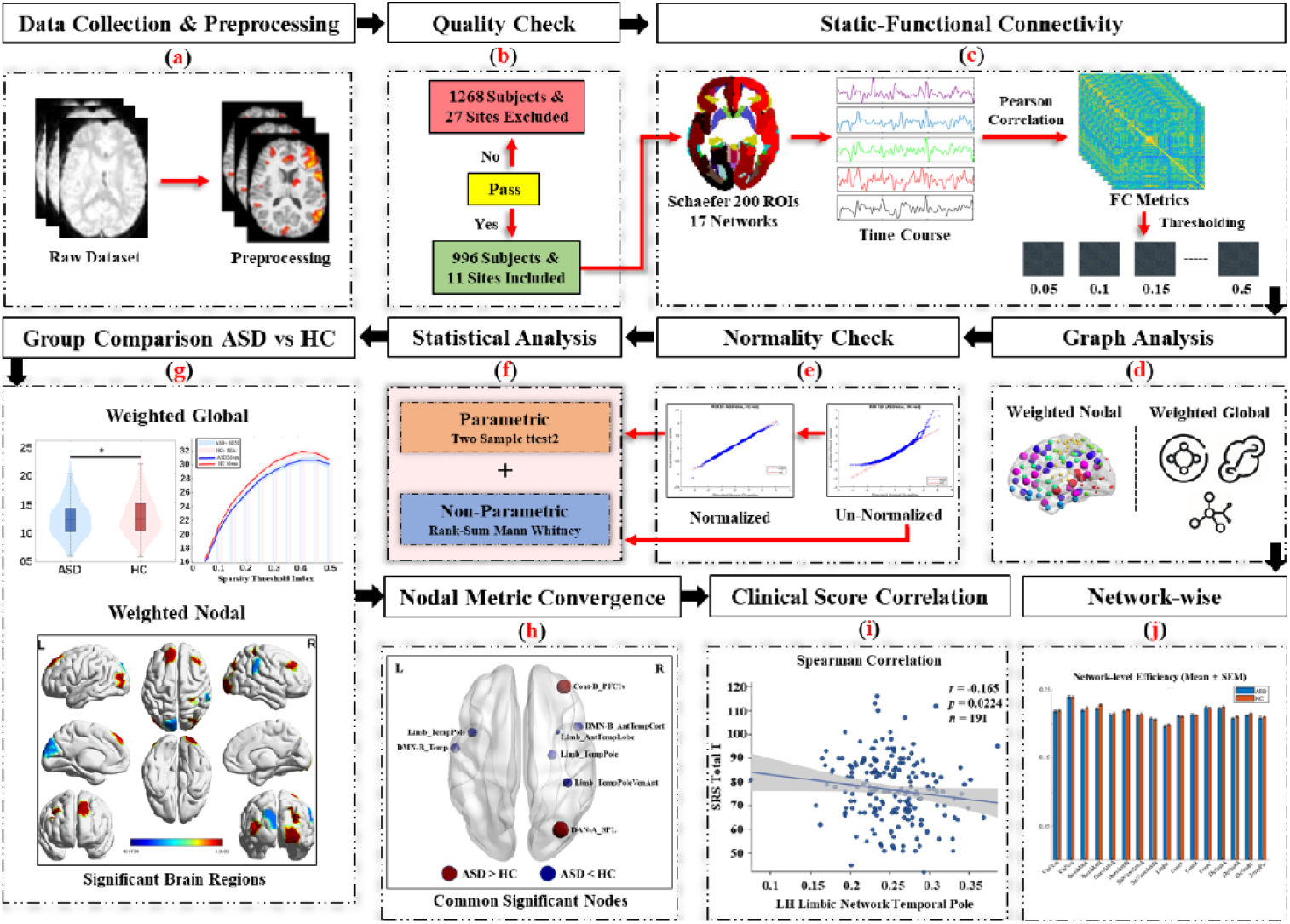
Overview of the schematic Pipeline processed method including (a) rs-fMRI data acquisition and preprocessing. (b) Quality check (c) Functional Connectivity (d) Graph Theoretical Construction (e) Normality check (f) Statistical analysis (g) Group Comparison (h) Nodal metric convergence (i) Clinical Correlation (j) System-wise analysis

The detailed research methodology employed in this study is outlined in Figure 1. The pipeline includes data acquisition of multi-site resting state functional magnetic resonance image from the ABIDE-I and ABIDE-II data repositories. After that preprocessing was done, following preprocessing the data went into quality check and filtered out sites and subjects. Functional connectivity was done using cleaned data. The rest of the steps including network construction, computation of nodal and global graph theoretical metrics and AUC based statistical analysis. This diagram is showing the key schematic process from collecting the raw data to the final network level results. This pipeline guarantees methodological reproducibility and transparency.

### 3.2 Global and Nodal Graph Theoretical Network Alterations

#### 3.2.1 Global Network Topology

Group differences in global network organization were evaluated using AUC values derived across the full sparsity range (0.05–0.50) for weighted functional brain networks. The assessed metrics encompassed assortativity, global efficiency, local efficiency, hierarchy, synchronization, and small-world properties. Two-sample *t*-tests were applied to AUC values to compare the ASD and HC groups. As illustrated in Figure 2A, significant group differences emerged for assortativity, local efficiency, and normalized characteristic path length (λ). Specifically, the ASD group exhibited significantly lower assortativity (ASD: 12.10 vs. HC: 12.54; *t* = −2.02, *p* = 0.043, Cohen’s *d* = −0.129), reduced local efficiency (ASD: 0.34 vs. HC: 0.35; *t* = −2.07, *p* = 0.038, Cohen’s *d* = −0.132), and decreased λ (ASD: 0.47 vs. HC: 0.48; *t* = −2.06, *p* = 0.039, Cohen’s *d* = −0.133), indicating subtle alterations in global integration relative to random networks organization, rather than changes in network segregation. No significant differences were observed for global efficiency, hierarchy, synchronization, or other small-world parameters (e.g., *Cp, Lp*, Gamma, Sigma). Although effect sizes were modest, such small effect sizes are typical of large-scale, multi-site neuroimaging studies and likely reflect distributed rather than focal network alterations. False discovery rate (FDR) correction was successfully applied to all reported global level results across tested global metrics (q < 0.05). Figure 2B illustrates threshold-wise group-averaged trajectories (mean ± SEM) across sparsity levels, demonstrating consistency of effects. The detailed statistical outputs are shown in Supplementary Table S3.

**Figure 2.**
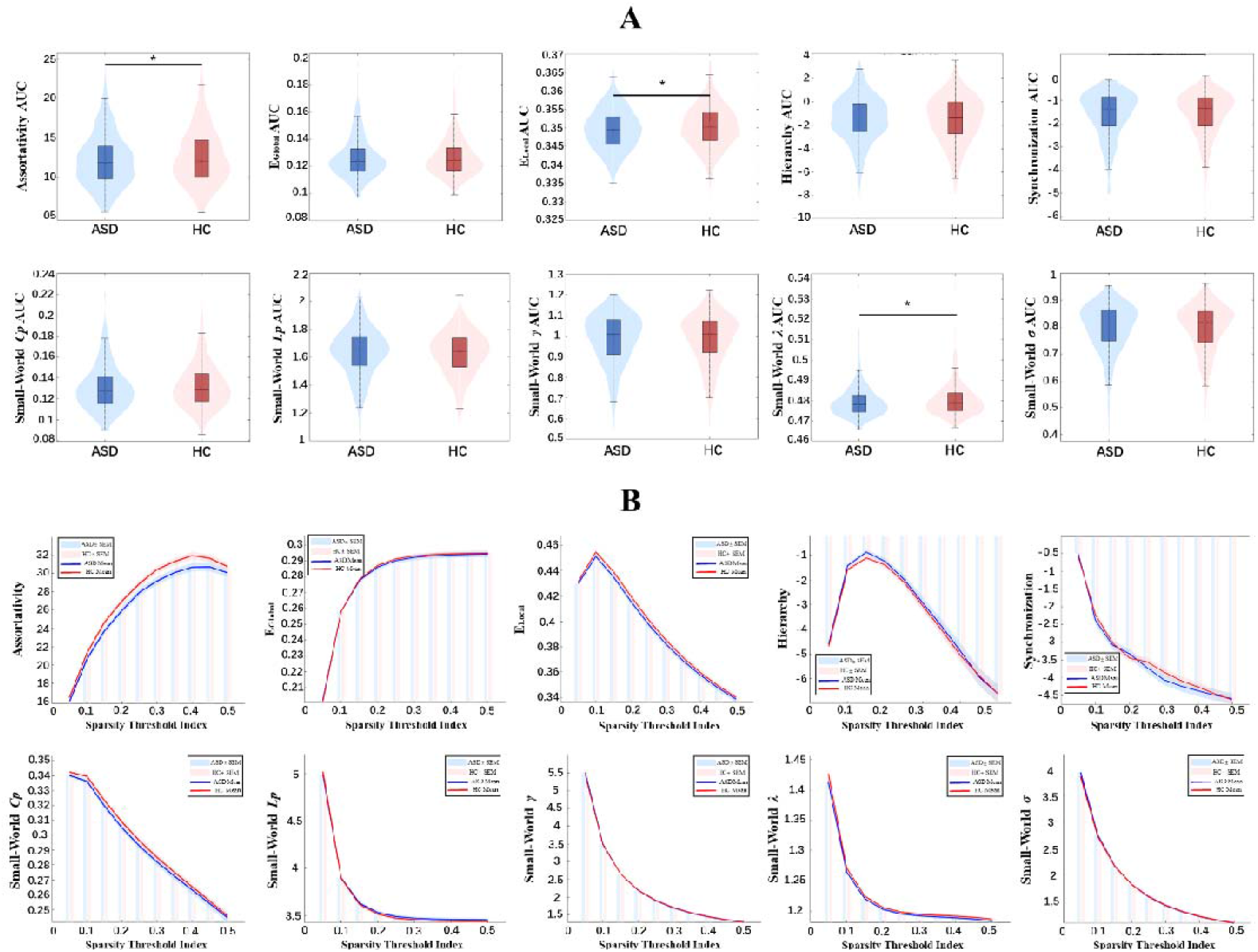
**(A)** AUC-based global graph-theoretical network alterations in ASD and HC **(B)** Threshold-wise comparison of global normalized assortativity, efficiency global, efficiency local, hierarchy, synchronization, small-worldness Cp, Lp, gamma, lambda, and sigma across sparsity levels. Mean ± SEM of z-scored all graph index values are shown for ASD and HC

#### 3.2.2 Nodal Network Topology

Significant group effects were identified for betweenness centrality, degree centrality, nodal efficiency, and shortest path length, while clustering coefficient and nodal local efficiency did not survive correction for multiple comparisons. Critically, the anatomical clustering of nodal effects within specific functional systems, rather than uniform distribution across the cortex, supports a system-level organization of nodal alterations in ASD. The most pronounced effects were localized to the default mode network (DMN), frontoparietal control network (FPN), dorsal attention network (DAN), limbic network, and Somatomotor network. Within these systems, the direction of change varied by region and metric, some areas showed increased centrality or efficiency in ASD, while others exhibited reductions. A complete listing of all regions with significant AUC-based nodal differences, including their network affiliations, statistical values, FDR-corrected *p*-values, effect sizes, and directional effects, is presented in Supplementary Table S4. Threshold-specific nodal results across the full sparsity range are reported in Supplementary Figures S1–S6 and Supplementary Tables S5–S10. Figure 3 displays the spatial distribution of AUC-based nodal differences between the ASD and HC groups.

**Figure 3.**
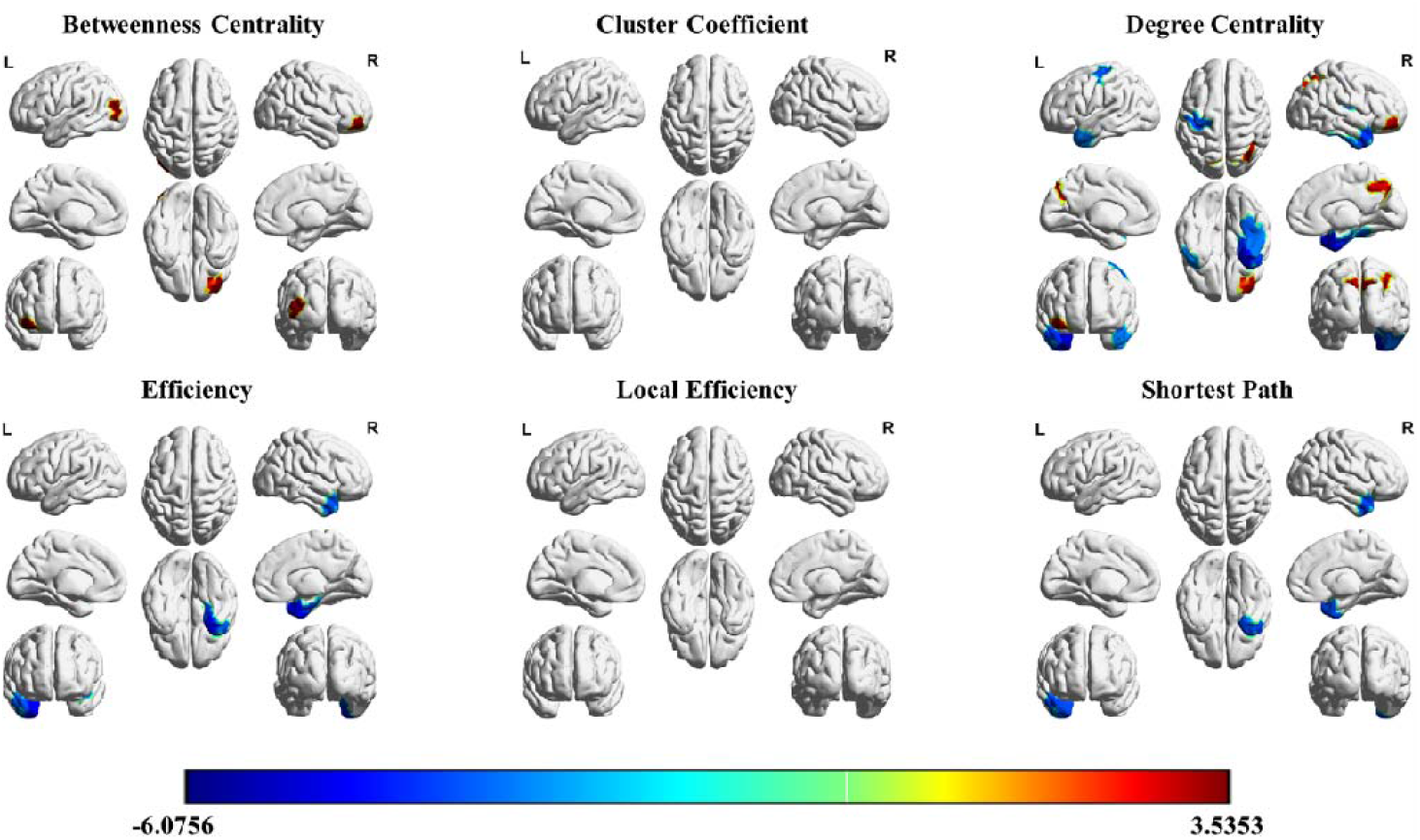
Spatial distribution of all brain regions showing significant AUC-based topological difference between ASD and HC

#### 3.2.3 Convergent Nodal Alterations Across Metrics

To distinguish robust and replicable network alterations from isolated or metric-specific findings, we identified regions showing consistent effects across two or more nodal graph metrics. Regions demonstrating increased nodal centrality or hub-like properties in the ASD group (ASD > HC) were primarily located in the right dorsal attention network, specifically the superior parietal lobule, as well as the right frontoparietal control network (encompassing the ventrolateral prefrontal cortex and inferior frontal gyrus), and midline DMN regions including the posterior cingulate cortex and precuneus. Conversely, regions showing reduced nodal efficiency and centrality in ASD (ASD < HC) were concentrated within the limbic network, particularly the bilateral temporal poles and anterior temporal cortices, as well as a DMN subnetwork (subnetwork B) linked to socio-emotional processing. Notably, several of these regions exhibited consistent directional effects across multiple complementary metrics, such as degree centrality, betweenness centrality, nodal efficiency, and shortest path length, suggesting a coherent, system-level reorganization of functional architecture in ASD rather than spurious or isolated statistical anomalies. All these convergent alterations are summarized in Table 4 and can be visualized in Figure 4.

**Figure 4.**
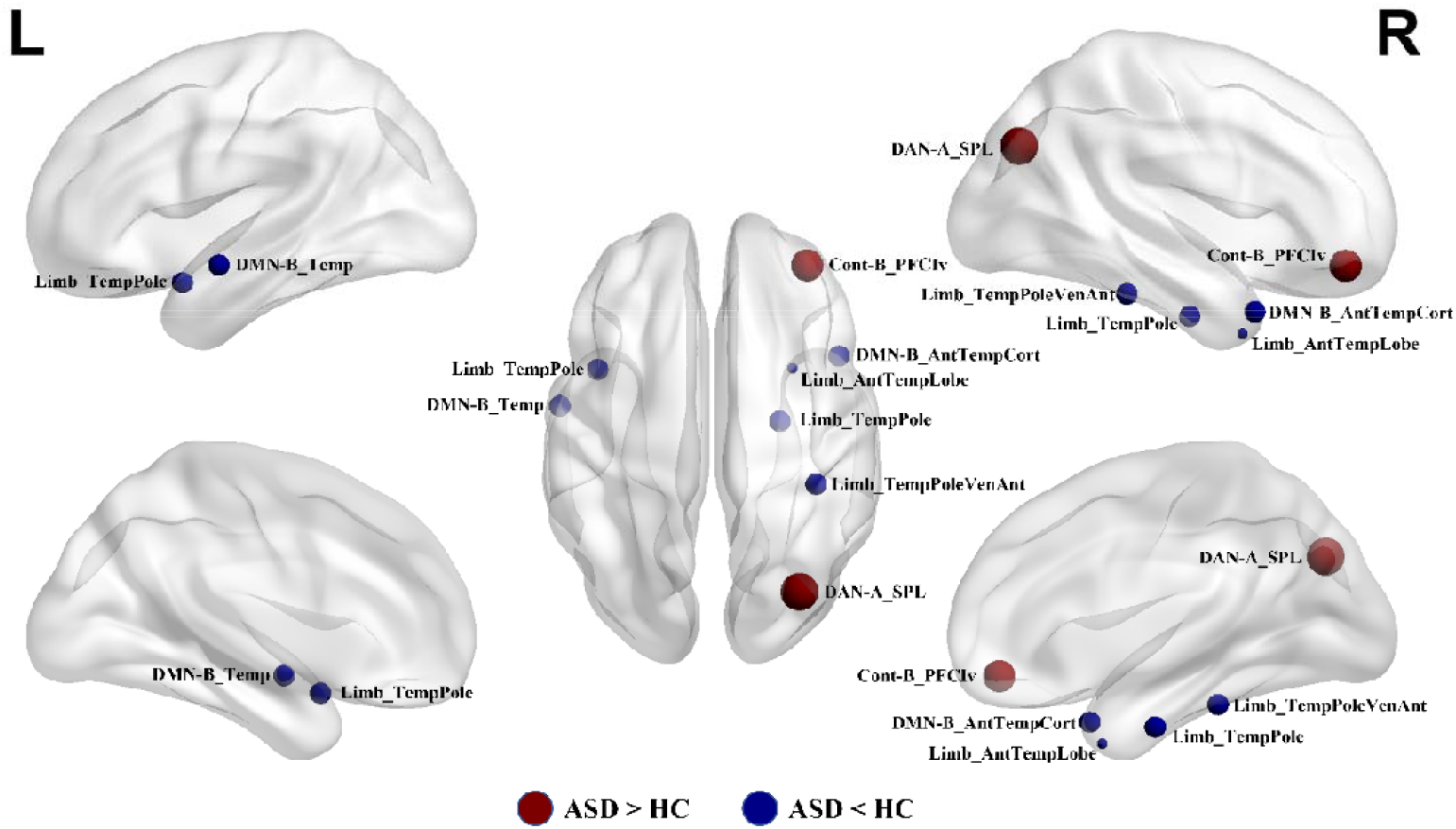
Nodes show regions with consistent group differences across multiple graph metrics using directional patterns, showing increased and decreased in ASD relative to HC. Increased nodal centrality or hub-like properties in ASD (ASD > HC) with red warm color and decreased nodal efficiency or centrality in ASD (ASD < HC) with blue warm color. Node size represents effect size (Cohen’s d). Only regions with convergent directional effects across multiple nodal metrics are shown.

**Table 3:** Common brain regions showing consistent area under the curve (AUC)-based nodal topological alterations across multiple graph-theoretical metrics, reflecting convergent changes in nodal centrality, efficiency, and network integration in ASD relative to healthy controls.

| Brain Regions | Hemisphere | Network | Metrics Showing Significance | *t*-value | *p*(*q*) | Direction |
| --- | --- | --- | --- | --- | --- | --- |
| Temporal Region | LH | Default mode network (DMN) (subnetwork-B) | Degree Centrality, Betweenness Centrality | -3.44 | 0.029 | ASD < HC |
| Superior Parietal Lobule (SPL) | RH | Dorsal Attention Network (DAN) (subnetwork-A) | Degree Centrality, Betweenness Centrality | 4.49 | 0.0013 | ASD > HC |
| Temporal Pole (BA38) | LH | Limbic | Degree Centrality, Nodal Efficiency | -3.73 | 0.007 | ASD < HC |
| Temporal Pole<br>(anterior<br>temporal lobe) | RH | Limbic | Degree Centrality,<br>Nodal Efficiency,<br>Shortest Path<br>Length | -3.06 | 0.029 | ASD <<br>HC |
| Temporal Pole | RH | Limbic | Degree Centrality,<br>Nodal Efficiency | -4.18 | 0.002 | ASD <<br>HC |
| Temporal Pole<br>(ventral/anterior) | RH | Limbic | Degree Centrality,<br>Betweenness<br>Centrality | -3.49 | 0.011 | ASD <<br>HC |
| **Ventrolateral**<br>**Prefrontal**<br>**Cortex (Inferior**<br>**Frontal Gyrus**<br>**region)** | **RH** | **Frontoparietal**<br>**Control**<br>**Network (FPN)**<br>**(subnetwork-B**<br>**)** | **Degree**<br>**Centrality,**<br>**Betweenness**<br>**Centrality** | **4.43** | **0.0009** | **ASD >**<br>**HC** |
| Anterior<br>Temporal Cortex<br>(ATC) | RH | Default Mode<br>Network<br>(DMN)<br>(subnetwork-B | Degree Centrality,<br>Nodal Efficiency,<br>Shortest Path<br>Length | -4.22 | 0.002 | ASD <<br>HC |

### 3.3 System-Level (Network-Wise) Topological Alterations

System-level analyses were performed by aggregating nodal graph metrics within each of the 17 Yeo functional networks and integrating results across sparsity thresholds using AUC summarization. Group differences between ASD and HC participants were evaluated using ANCOVA models, with age, sex, and acquisition site included as covariates to account for potential confounding effects. Significant between-group differences emerged across multiple functional systems, with the most pronounced effects observed in higher-order associative networks, particularly the default mode, frontoparietal control, and limbic networks. Alterations were especially evident, including degree centrality, clustering coefficient, and nodal efficiency, suggesting a systematic reconfiguration of large-scale functional architecture in ASD. Group differences between ASD and healthy controls (HC) in network level graph metrics with adjusted covariates of AUC-based metrics can be visualized in Figure 5. The stability of these system-level findings across the full range of sparsity thresholds is demonstrated in Supplementary Figure S7, while inter-subject variability and distributional characteristics of network metrics are illustrated in Supplementary Figure S8. Comprehensive MANCOVA results, including F-statistics, uncorrected *p*-values, and FDR-corrected *p*-values, for each network and metric are provided in Supplementary Tables S11–S16.

**Figure 5.**
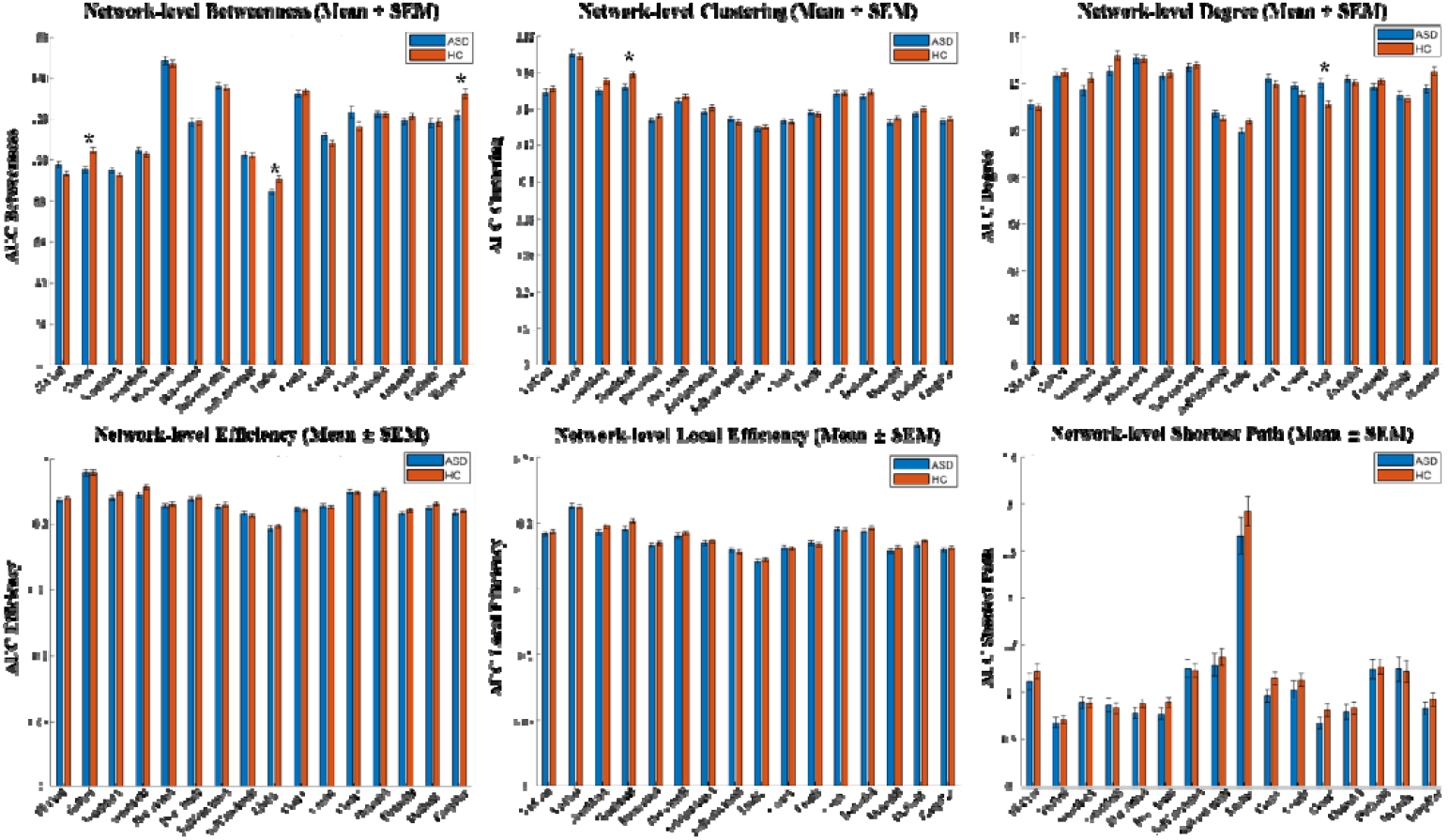
Covariate-adjusted mean ± SEM of area-under-the-curve (AUC)–based network-level graph metrics are shown for the ASD and healthy control (HC) groups across the 17 Yeo functional networks. Group differences were assessed using ANCOVA with age, sex, and acquisition site included as covariates.

### 3.4 Clinical–Graph Correlation Analysis

To evaluate the clinical relevance of observed topological alterations, we conducted Spearman rank correlation analyses between nodal graph-theoretical metrics and standardized measures of ASD symptom severity. These analyses were restricted to a targeted set of regions. This targeted approach was adopted to reduce multiple-testing burden and focus on regions showing convergent, robust group-level effects. The regions included were the left temporal region of DMN subnetwork B, the left temporal pole within the limbic network, the right superior parietal lobule in the dorsal attention network, and the right anterior temporal pole (limbic network).

Nodal topology in the left DMN temporal region showed significant negative correlations with both ADOS-G Total score (*r* = −0.179, *p* = 0.0017, *df* = 302) and ADOS-2 Severity Total (*r* = −0.129, *p* = 0.0356, *df* = 266), indicating that lower nodal integration in this region was associated with greater symptom severity. Similarly, the left limbic temporal pole exhibited negative correlations with SRS Total T score (*r* = −0.165, *p* = 0.0224, *df* = 191) and ADI-R Social Total A (*r* = −0.125, *p* = 0.0257, *df* = 318). In contrast, the right superior parietal lobule (dorsal attention network) a region showing increased centrality in ASD, demonstrated a positive correlation with ADI-R Social Total A (*r* = 0.130, *p* = 0.0203, *df* = 318. A smaller but significant positive association was also observed between ADI-R Social Total A and nodal topology in the right limbic temporal pole (*r* = 0.111, *p* = 0.048, *df* = 318). Although correlation magnitudes were modest, the directional consistency between group-level network alterations and clinical associations, particularly within the default mode, limbic, and attention systems, supports a modest but consistent association between intrinsic brain network organization and core behavioral features of ASD. All the clinical correlation results are illustrated in Figure 6.

**Figure 6.**
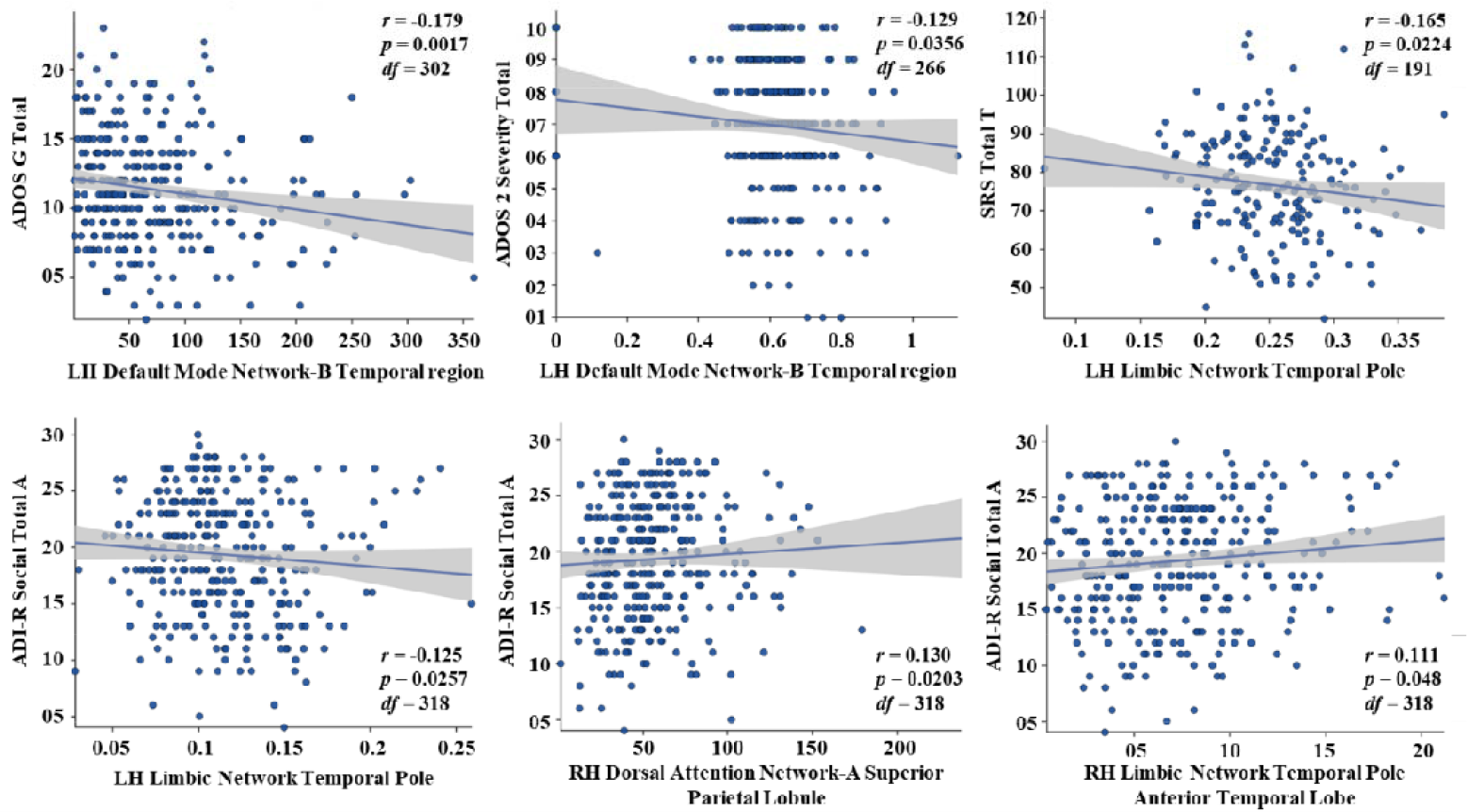
Scatter plots illustrate Spearman rank correlations between AUC-based nodal graph-theoretical metrics and standardized clinical measures of ASD symptom severity (ADOS-G, ADOS-2, SRS, and ADI-R). Analyses were restricted to regions showing convergent group-level alterations, including default mode, limbic, and dorsal attention network nodes. Each point represents an individual participant, with correlation coefficients (*r*) and corresponding *p*-values indicated.

## 4 Discussion

The present research identified significant alterations in global functional network architecture in ASD participants by showing decreased assortativity, local efficiency and normalized characteristics path lengths (lambda) [71]. Lower assortativity suggests altered degree–degree organization, potentially reflecting reduced preference for connections among similarly connected nodes in ASD networks [56]. These alterations may indicate reduced tolerance to perturbations within local subnetworks[72]. Reduced normalized path length (λ) suggests subtle deviations in small-world balance relative to random network organization[12]. The important thing is that the results were similar across AUC-based analyses, indicating stability with a relatively small effect size. Although effect sizes were small, this pattern is consistent with prior large-scale ASD neuroimaging studies and likely reflects distributed network alterations rather than focal effects.

At the nodal level, the changes were not randomly distributed throughout the brain related with autism, but rather showed significant clustering in advanced order associative network [73]. The increase in nodal betweenness and centrality were observed in the brain regions that are associated with the default mode network, frontoparietal control network, and dorsal attention network. The brain regions include posterior cingulate cortex, precuneus, superior parietal lobule, and ventrolateral prefrontal cortex. These regions are strongly implicated in attentional control, self-referential processing, and executive function [74]. Compared to that, lower nodal efficiency and centrality were noticed mostly in the limbic and anterior temporal regions, specifically the temporal pole, which is important for socio-emotional processing and social cognition [75]. The convergence of alterations across different nodal graph metrics within the same regions indicates strong topological configurations rather than metric-specific effects. This supports the presence of systematic network-level reorganization in ASD.

System-level investigation additional revealed that ASD related topological alterations affect whole functional systems, instead of just individual nodes [76]. Significant changes were found throughout different Yeo defined networks, particularly in high order associative systems including the default mode, attention and control networks [77]. These results were consistent with hypotheses that suggest altered hierarchical organization of large-scale brain networks in ASD. Significantly, the concentration of alterations within high order associative systems shows that the ASD related network reorganization is most effectively understood at the level of integrated functional systems rather than isolated brain regions. The consistency of system-level group differences throughout a broad range of network sparsity thresholds, along with statistical control for age, sex and acquisition sites, suggests that these effects reflect robust alterations in large scale functional network organization instead of threshold-specific or site driven artifacts.

In addition, the clinical graph correlation research identified significant connections between altered nodal topology and central ASD symptom severity. The decrease in nodal centrality and efficiency in limbic and default mode network were associated with increased social and communication impairments indicated by ADOS, SRS and ADI-R scores [78, 79]. However, increased in nodal centralities in dorsal attention regions was positively linked to social symptom severity and may reflect altered engagement of attentional control systems associated with symptom severity [80, 81]. The potential clinical significance of large-scale functional network topology in ASD can be seen by the identified clinical graph correlations. In addition, higher social and communication impairments measured by the ADOS, ADI-R, and SRS measures were slightly associated with decreased nodal centrality and efficiency within limbic and default mode regions. On the other hand, there were positive correlations between the symptom severity and increased nodal centrality within dorsal attention regions. This may reflect because of altered or compensatory engagement of attentional control systems. The consistency with group level network alterations indicates that individual differences in intrinsic network organization may be strongly related to major ASD symptom aspects, even though, these associations were small in magnitude and do not imply causality.

Several limitations should be acknowledged. First, despite rigorous preprocessing and quality control procedures, multi-site resting-state fMRI data inherently involve variability arising from differences in scanner hardware, acquisition protocols, and site-specific factors. Although site effects were statistically controlled, residual confounding cannot be entirely excluded. Second, the cross-sectional design limits inferences regarding developmental trajectories and causal relationships underlying network alterations in ASD. Third, the observed effect sizes were modest, which is typical for large-scale neuroimaging studies but nonetheless warrants cautious interpretation. Finally, while the area-under-the-curve framework enhances robustness across network thresholds, graph-theoretical metrics remain sensitive to methodological choices, including parcellation strategy and connectivity construction.

Future studies should prioritize longitudinal designs to characterize developmental changes in functional network topology across the lifespan in ASD. Integrating multimodal neuroimaging data, such as structural MRI and diffusion-weighted imaging, may further clarify the neurobiological substrates underlying observed topological alterations. In addition, combining graph-theoretical features with machine learning approaches could improve individual-level prediction of symptom severity and treatment response. Extending this work to task-based fMRI and dynamic functional connectivity analyses may also provide complementary insights into state-dependent network reconfiguration in ASD.

## Conclusion

In conclusion, this study provides a multi-scale characterization of functional brain network topology in autism spectrum disorder using a large, multi-site resting-state fMRI dataset. By integrating global, nodal, and system-level graph-theoretical measures with clinical correlation analyses within an area-under-the-curve framework, the findings indicate that ASD is associated with organized, non-random reconfiguration of large-scale functional brain networks, rather than uniform or focal disruption. Topological alterations were most consistently observed within higher-order associative systems, including the default mode, limbic, dorsal attention, and frontoparietal control networks, reflecting an altered balance between network integration, segregation, and hub organization. Associations between nodal topology and clinical symptom severity further support the functional relevance of these network-level changes. Together with complementary component-level connectivity findings, the present results support a convergent framework in which ASD is best understood as a disorder of distributed brain network organization rather than isolated regional dysfunction. By leveraging robust graph-theoretical methodology and a large heterogeneous cohort, this work advances a reproducible, systems-level understanding of ASD and provides a foundation for future longitudinal and multimodal investigations.

## Acknowledgement

The authors thank the Brain Connectivity Lab and University of Electronic Science and Technology of China for technical support. We are grateful to the developers of the ABIDE dataset and the neuroimaging community for making the data publicly available. We also acknowledge the assistance of colleagues who helped review earlier versions of the manuscript.

## Data Availability Statement

The resting-state fMRI data used in this study are publicly available from the Autism Brain Imaging Data Exchange (ABIDE) repository at http://fcon_1000.projects.nitrc.org/indi/abide/. This dataset includes contributions from 38 different sites worldwide, encompassing a total of 2,264 participants, with 1,083 subjects diagnosed with ASD and 1,181 healthy controls.

## CRediT Author Contribution Statement

**Talha Imtiaz Baig:** Conceptualization, Methodology, Visualization, Writing – original draft. **Hongzhou Wu:** Conceptualization, Methodology, Investigation. **XiYang Li:** Visualization, Software. **Junlin Jing:** Data curation, Formal analysis. **Bharat B. Biswal:** Supervision, Validation, Investigation, Writing – review & editing. **Benjamin Klugah-Brown:** Funding acquisition, Validation, Writing – review & editing, Supervision, Resources.

## Supporting Information

Additional supporting information can be found online in the Supporting Information section (Supplementary Results/Methods).

